# Obstetric Referral Practices and Health System Factors in Public Health Centres of Addis Ababa, Ethiopia: A Mixed-Methods Study

**DOI:** 10.64898/2026.01.31.26345258

**Authors:** Amaha Haile Abebe, Rose Mmusi-Phetoe

## Abstract

**Objective:** To determine the magnitude of obstetric referral and to explore contextual factors influencing referral practices in public health centres of Addis Ababa, Ethiopia.

**Design:** Explanatory sequential mixed-methods study.

**Setting and period:** Fifty public health centres in Addis Ababa City Administration, January–April 2021.

**Methods:** Delivery and referral registers from 50 health centres were reviewed retrospectively for 12 months (8 July 2019–7 June 2020). Facility observations and interviews with maternity unit heads were conducted in all selected centres. In-depth interviews were conducted with 20 midwives and 13 health-centre managers. Quantitative data were analysed descriptively, and qualitative data were analysed thematically using Colaizzi’s method.

**Results:** Eighty percent of health centres had a functional referral system. The overall obstetric referral rate was 32%, with substantially higher referral rates in facilities without caesarean section (CS) services compared with those providing CS (39% vs 21%). Qualitative findings indicated that high referral rates were associated with limitations in the predictive capacity of the partograph, variability in providers’ clinical skills, and risk-averse practices driven by accountability concerns related to maternal and perinatal outcomes.

**Conclusion:** Although referral systems were largely functional, obstetric referral rates were high, suggesting potential over-referral. Updating labour monitoring tools, strengthening provider competencies, and clarifying accountability mechanisms may reduce unnecessary referrals.

## Introduction

Obstetric complications are often unpredictable and can rapidly progress to severe, life-threatening conditions. Consequently, women who develop complications at lower levels of care must be promptly referred to facilities capable of providing comprehensive emergency obstetric care, including caesarean section delivery [1]. The World Health Organization (WHO) quality-of-care framework identifies a functional referral system as one of the eight core domains of quality maternal and newborn care [2].

The WHO quality standards emphasize that every woman and newborn with conditions beyond the capacity of a facility should be appropriately and promptly referred. These standards underscore timely assessment at admission, during labour, and in the early postnatal period; rapid decision-making; pre-established referral plans; and effective communication and feedback between referring and receiving facilities [2].

Effective referral systems are essential for reducing delays in accessing appropriate care. However, referral capacity at the primary care level remains weak in many sub-Saharan African countries. Analysis of service availability data from five countries showed that only 38% of primary care facilities and 78% of secondary care facilities had adequate referral capacity[3].

Reliable transport—typically ambulances available 24 hours a day—is a critical component of effective referral systems [2]. In Ethiopia, only 17% of health facilities have a dedicated, functioning ambulance, with many relying on shared district-level services. Approximately one-third of facilities lack any ambulance service, requiring women to arrange their own transport[4]. Similar challenges have been reported in Malawi[5].

Beyond availability, ambulance management—including fuel supply, driver availability, and prioritization for emergencies—directly affects referral timeliness. In Indonesia, referral delays persisted despite universal ambulance availability due to administrative and staffing constraints[6].

Effective referral systems also depend on structured communication and feedback mechanisms. Although most Ethiopian facilities assign staff to accompany referred women, advance notification and systematic feedback from referral hospitals remain uncommon [4]. Comparable gaps have been documented in South Africa[7].

Referral decisions must be timely and appropriate. Unnecessary referrals burden higher-level facilities and impose avoidable costs on families. The WHO estimates that 10–15% of pregnant women develop complications requiring caesarean section [8]. Primary health care facilities without CS capability would therefore be expected to refer approximately this proportion. However, reported referral rates often exceed this range, from 10–63% in the Netherlands [9] to 28% in Tanzania [10].

In Ethiopia, 73% of women with obstetric complications admitted to health centres were referred, suggesting limited capacity or underutilization of available services [4]. Against this background, this study assessed the magnitude and contextual drivers of obstetric referral in public health centres in Addis Ababa City.

## Research methods

### Study design and setting

An explanatory sequential mixed-methods design was used. A quantitative phase involving retrospective record review was followed by a qualitative phase using in-depth interviews to explain quantitative findings. The quantitative and qualitative data collection was started on January 1, 2021, and completed on April 28, 2021. The study was conducted in 50 public health centres of the Addis Ababa City Administration.

### Quantitative component

#### Sampling and data sources

Fifty public health centres were randomly selected from the 90 centres providing delivery services in Addis Ababa. Five centres were selected from each of the city’s ten sub-cities using a lottery method. Delivery, referral, and operating theatre registers were reviewed during the above-mentioned study period. Record review covered registers for the Ethiopian Fiscal Year 2012 (8 July 2019–7 June 2020). Facility interviews and observations using a structured checklist assessed the functionality of referral systems. The authors did not collect or access data that identifies individual clients’ identities, either during or after the data collection

#### Data collection

Trained midwives extracted data using a structured checklist capturing service volume, mode of delivery, operative vaginal delivery, referrals, and caesarean section procedures. Interviews and observations were conducted using a structured questionnaire to assess the availability of the functional referral system. Supervisors conducted daily data quality checks.

#### Data analysis

Quantitative data were entered into EpiData and analysed using SPSS version 20. Descriptive statistics (means, medians, proportions, ranges) were used. Obstetric referral rates were calculated for facilities for the Ethiopian Fiscal Year 2012 (8 July 2019–7 June 2020). Referral rate was calculated by dividing the annual number of women referred for obstetric and newborn care-related cases to higher level facilities from the health centres by the total number of obstetric and newborn care cases served (delivered in the same health facility + referred to higher level care) by the health centers multiplied by 100.

### Qualitative component

#### Sampling and data collection

Purposive sampling identified 20 midwives and 13 health centre managers directly involved in maternity care and service management. In-depth interviews were conducted using a semi-structured guide, audio-recorded with consent, and supplemented by field notes.

#### Data analysis

Interviews were transcribed verbatim and analysed thematically using Colaizzi’s seven-step method to identify themes related to referral decision-making and practice.

### Ethical considerations

Ethical approval was obtained from the Research Ethics Committee of the Department of Health Studies, University of South Africa, and the Addis Ababa City Administration Health Bureau.

Written informed consent was obtained from all participants, and confidentiality was maintained.

## Results

### Functionality of referral systems

Overall, 80% (n=40) of health centres met criteria for a functional referral system (≥6 of 8 indicators). Most centres had a functioning ambulance (84%) and reliable communication mechanisms (92%). While standardized referral forms and documentation were widely available, only 36% of centres reported receiving timely, case-by-case feedback from referral hospitals (Table-1).

**Table 1:** Percentage distribution of health centres with functional referal system (N=50)

| Functioning Referral System | No (%) | Yes (%) |
| --- | --- | --- |
| Owned a functioning ambulance for the emergency transport of women and newborns to referral facilities | 16 | 84 |
| Had a functioning communication method (a mobile phone, land line or radio) for referrals and consultation on complicated cases | 8 | 92 |
| Had an up-to-date list of network facilities that provide referral obstetric and newborn care | 4 | 96 |
| Had formal agreements, communication arrangements, and a feedback system with referral centre(s) | 34 | 66 |
| Had a standardized referral form that enabled them to document relevant clinical information and the reason for referral | 4 | 96 |
| Had appropriate documentation or register for referred cases | 10 | 90 |
| Always assigned professionals to accompany cases to referral facilities | 10 | 90 |
| Regularly received case-by-case timely feedback | 64 | 36 |
| Had effective referral system (scored $\geq 6$ out of 8 points) | 20 | 80 |

### Obstetric Referral rates

The overall obstetric referral rate was 32%. Referral rates were higher in facilities without CS services (39%) than in those providing CS (21%). Only five facilities reported referral rates below the expected 15% threshold.(Table-2).

**Table 2:**
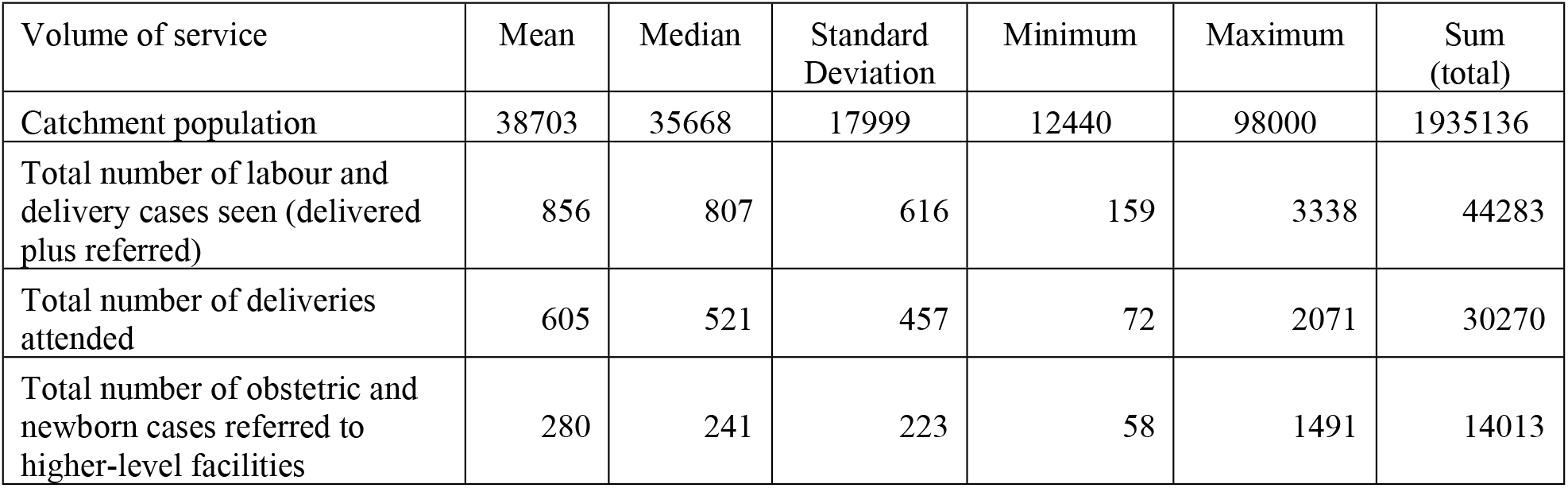
The mean number of women provided labour and delivery care and obstetric referals during Ethiopian Fiscal Year (EFY) 2012 (8 July 2019 –7 June 2020) (N=50)

| Volume of service | Mean | Median | Standard Deviation | Minimum | Maximum | Sum (total) |
| --- | --- | --- | --- | --- | --- | --- |
| Catchment population | 38703 | 35668 | 17999 | 12440 | 98000 | 1935136 |
| Total number of labour and delivery cases seen (delivered plus referred) | 856 | 807 | 616 | 159 | 3338 | 44283 |
| Total number of deliveries attended | 605 | 521 | 457 | 72 | 2071 | 30270 |
| Total number of obstetric and newborn cases referred to higher-level facilities | 280 | 241 | 223 | 58 | 1491 | 14013 |

### Qualitative Findings

Four interrelated themes emerged: availability and functionality of referral systems; limitations of the partograph; variability in providers’ clinical skills; and risk-averse practices driven by accountability pressures.

Participants reported that referral tools and ambulances were generally available. Despite widespread ambulance availability, operational challenges—including shared ambulance use, fuel shortages, and driver availability—undermined timely referral in some facilities. In addition, all noted the absence of timely feedback from referral hospitals.

Referral rates were widely perceived as high. Midwives attributed unnecessary referrals to the limited predictive value of the partograph, noting that many women crossed the action line but subsequently delivered normally at referral hospitals: A midwife said

> *“Most women cross the action line, yet they deliver normally at the hospital. The partograph needs to be improved for its ability to detect real problems.”*

Variability in clinical skills, particularly cervical dilatation assessment, was also identified. Managers emphasized risk aversion driven by strict accountability for maternal and perinatal deaths, which encouraged early referrals.: A manager said

> *“Providers prefer to refer early because of strict accountability for maternal and perinatal deaths.”*

## Discussion

This study demonstrates that public health centres in Addis Ababa largely possess the structural components of functional obstetric referral systems. However, high obstetric referral rates indicate that infrastructure availability alone does not ensure appropriate and efficient referral practice. The findings highlight a critical gap between system readiness and actual referral performance.

Despite widespread ambulance availability, operational challenges—including shared ambulance use, fuel shortages, and driver availability—undermined timely referral in some facilities. These findings are consistent with evidence from other low- and middle-income settings, where referral delays persist due to governance and management constraints rather than lack of physical resources [5, 6].Strengthening emergency transport governance is therefore essential to improving referral effectiveness.

Communication mechanisms and standardized referral documentation were largely in place, consistent with WHO recommendation [2]. However, weak feedback and counter-referral mechanisms limited opportunities for clinical learning and quality improvement. The absence of systematic feedback may reinforce defensive referral practices, as providers are unable to evaluate referral appropriateness or outcomes. Similar challenges have been reported in South Africa [7].

Referral rates observed in this study substantially exceeded WHO estimates of the expected number of obstetric complications requiring referral or caesarean section [8]. Qualitative findings suggest that outdated labour monitoring tools, variability in provider skills, and risk-averse accountability environments contributed to over-referral. These findings align with WHO intrapartum care recommendations that question the predictive validity of the traditional partograph and support revised labour monitoring approaches [11, 12].

Strengthening provider competencies, updating labour monitoring tools, and implementing supportive accountability frameworks may help reduce unnecessary referrals while maintaining patient safety.

## Conclusion

Although referral systems in Addis Ababa public health centres are largely functional, obstetric referral rates are high, suggesting potential over-referral. Addressing this requires standardized referral guidelines, updated labour monitoring aligned with WHO recommendations, reliable emergency transport management, structured feedback mechanisms, and supportive accountability frameworks. These measures could improve efficiency and maternal and newborn outcomes.

## Data Availability

All relevant data are within the manuscript and its Supporting Information files.

## DECLARATION

### Ethics approval and consent to participate

The research protocol was reviewed and approved by the Research Ethics Committee of the Department of Health Studies of the University of South Africa. The research protocol was again reviewed and approved by the Ethical Review Committee of the Addis Ababa city administration health office. Once the research protocol had been approved by the ethical review committees, support letters were written from the Addis Ababa city administration health office and sub-city health offices to study health facilities.

Written informed consent was obtained from all study participants, and interviews were conducted in a setting that ensured privacy and confidentiality.

### Consent for publication

Not applicable. Our manuscript does not contain data from any individual person

### Availability of data and materials

The datasets used and/or analyzed during the current study are available from the corresponding author on reasonable request.

### Authors’ contributions

Amaha Haile Abebe, Corresponding author have conceptualized and designed the study protocol, coordinated, supervised, and conducted the data collection, data entry, analysis, and report write-up. Prepared the manuscript.

Prof. Rose Mmusi-Phetoe, co-author, supervised and contributed to conceptualizing and designing the study protocol, data analysis, and report write-up and review. She further reviewed and refined the manuscript.

## Acknowledgments

We would like to thank the University of South Africa for financing the study. We would like to thank women, midwives, and health center heads in Addis Ababa city for participating in the study. I would like to thank the research assistants who conducted the qualitative data collection, namely, Sr. Hawa Ali, Sr. Hasna Musema, Sr. Aselefech Negewo, and Sr. Abeba Gebrehiwot.

## Abbreviations

CS: Caesarean Section
SPSS: Statistical Package for the Social Sciences
UNISA: University of South Africa
WHO: World Health Organization

